# SARS-CoV-2 qRT-PCR Ct value distribution in Japan and possible utility of rapid antigen testing kit

**DOI:** 10.1101/2020.06.16.20131243

**Authors:** Yuta Takeda, Masatoshi Mori, Kazuya Omi

## Abstract

The exact pathology of COVID-19 remains mostly unclear, and accurate epidemiological understanding and rapid testing are crucial to overcome this disease. Several types of nucleic acid tests (NAT) have been used in Japan, but information about the viral RNA load, determined by Ct values, of the patients is limited due to the small number of patients tested in each clinical institution and lack of standardization of the testing kits. We have been performing the qRT-PCR tests established by NIID, and the mean Ct value distribution of 62 cases, which are deemed “first-visit” patients, among the total of 88 positive cases tested in a 4-day window of early April, was 24.9 with SD=5.45. Recently approved antigen testing kits were also used in the same samples (62 positives) along with 100 negative cases, and it revealed the positive predictive value of 80.6% and negative predictive value of 100%, with an overall agreement rate of 92.6%. These results indicate that a certain number of patients with lower Ct values, existed in Japan when SARS-CoV-2 virus started to spread. The newly approved rapid antigen testing kit will be a useful tool to identify such populations rapidly.

## Introduction

Coronavirus disease 2019 (Covid-19) pandemic is caused by the SARS-CoV2 virus, and the global number of cases has surpassed 6 million. In Japan, RT-PCR testing has been conducted primarily to high-risk patients, i.e., exhibiting persistent fever, chest pain, or who are elderly ^1,2,3^. The first wave coming from Wuhan was epidemic, and the strain brought from Europe and the US caused the pandemic in April 2020, affecting more than 15,000 people in the country by the end of April^4^. COVID-19 infection is diagnosed by the detection of SARS-CoV-2 RNA in nasopharyngeal specimens, and in Japan qRT-PCR method established by the National Institute of Infectious Diseases (NIID) has been widely used. However, the cycle threshold (Ct) value has not been standardized and the small number of patients tested in each clinical institution, making the interinstitutional comparison difficult. As far as we know, no reports have been made to describe Ct value distribution in Japan so far. Here we measured the Ct-value distribution in the samples tested in SRL during a 4-day period in early April, when the second wave by SARS-CoV-2 virus from EU hit Japan ^5^, using the N2 probe, which is the standardized NIID method. Recently, a SARS-CoV-2 rapid antigen detection kit has been developed and approved by Ministry of Health, Labour and Welfare. The rapid COVID-19 antigen testing kit, which was approved in Japan in 2020 May, is expected to be used to confirm positivity in new symptomatic patients, not to confirm negativity or to follow-up patients previously confirmed to be SARS-CoV-2 positive (https://www.mhlw.go.jp/content/10900000/000631469.pdf). The antigen detection kit is considered to show lower sensitivity compared to qRT-PCR, which is a definitive diagnostic method for COVID-19, and its clinical usefulness has not been established. The relationship between the CT-value distribution and the positivity of the rapid antigen detection kit was also investigated.

## Materials and Methods

Specimens are collected by each clinical institution in early April, and clinical testing for SARS-CoV-2 were performed by SRL Inc. upon request from institutions. In SRL Inc., each specimen is given a unique ID code upon receipt, and when specimens from an individual are collected multiple times (e.g., for follow-up purposes or to test if the patient meets hospital discharge criteria), they are labeled with the same ID as the initial receipt. In this report, we have defined the specimens received for the first time in SRL as “Initial ID samples”. In this survey, we have not received and used any personal identifiable information and clinical information.

RNA was extracted from nasopharyngeal swabs using QIAsymphony and Virus/Pathogen kit (Qiagen). qRT-PCR tests were performed as follows, according to the manual provided by NIID (https://www.niid.go.jp/niid/images/epi/corona/2019-nCoVmanual20200217-en.pdf). One step qRT-PCR was performed using a QuantiTect Probe RT-PCR Kit (Qiagen). 7500 Real-time PCR Systems (Thermo Fisher Scientific) were used and Ct-values were obtained using N2 primer (NIID_2019-nCOV_N_F2, NIID_2019-nCOV_N_R2) and probe (NIID_2019-nCOV_N_P2). Rapid antigen testing was carried out using ESPLINE SARS-CoV-2 (Fujirebio Inc). Nasopharyngeal swabs were mixed with the sample treatment solution and incubated for 5 min. 20 µL of the treated sample was applied to the kit and was incubated for 30 minutes. After incubation, test and reference lines were visually assessed by two staffs.

## Results

The total number of specimens tested for SARS-CoV-2 in the 4 days was 1425, and out of these, 88 (6.2%) were qRT-PCR positive. These positive specimens included 62 Initial ID nasopharyngeal swabs. The Ct value distribution of all positive specimens was 27.0 ± 6.25 (mean ± SD), Initial ID specimens was 24.9 ± 5.45 (mean ± SD) (Figure 1). The rapid antigen test kit is indicated to be used to confirm positivity in new symptomatic patients, not to confirm negativity or to follow-up patients previously confirmed to be SARS-CoV-2 positive. To evaluate its utility in our company and performance, we have tested all 62 Initial ID samples and randomly selected 100 qRT-PCR negative specimens (162 cases total) using the rapid antigen testing kit. Table 1 shows the positivity rate in each Ct value range. The positive rate of Ct values less than 25 was 100% (32/32). The positivity rates in Ct value range of 25 to 30 and 30 to 40 were 88.9% (16/18) and 16.7% (2/12) respectively. All the 100 qRT-PCR negative specimens were negative in the rapid antigen testing kit. The overall agreement rate was 92.6% (150/162), with positive predictive value (PPV) of 100% (50/50) and negative predictive value (NPV) of 89.3% (100/112).

**Figure 1.**
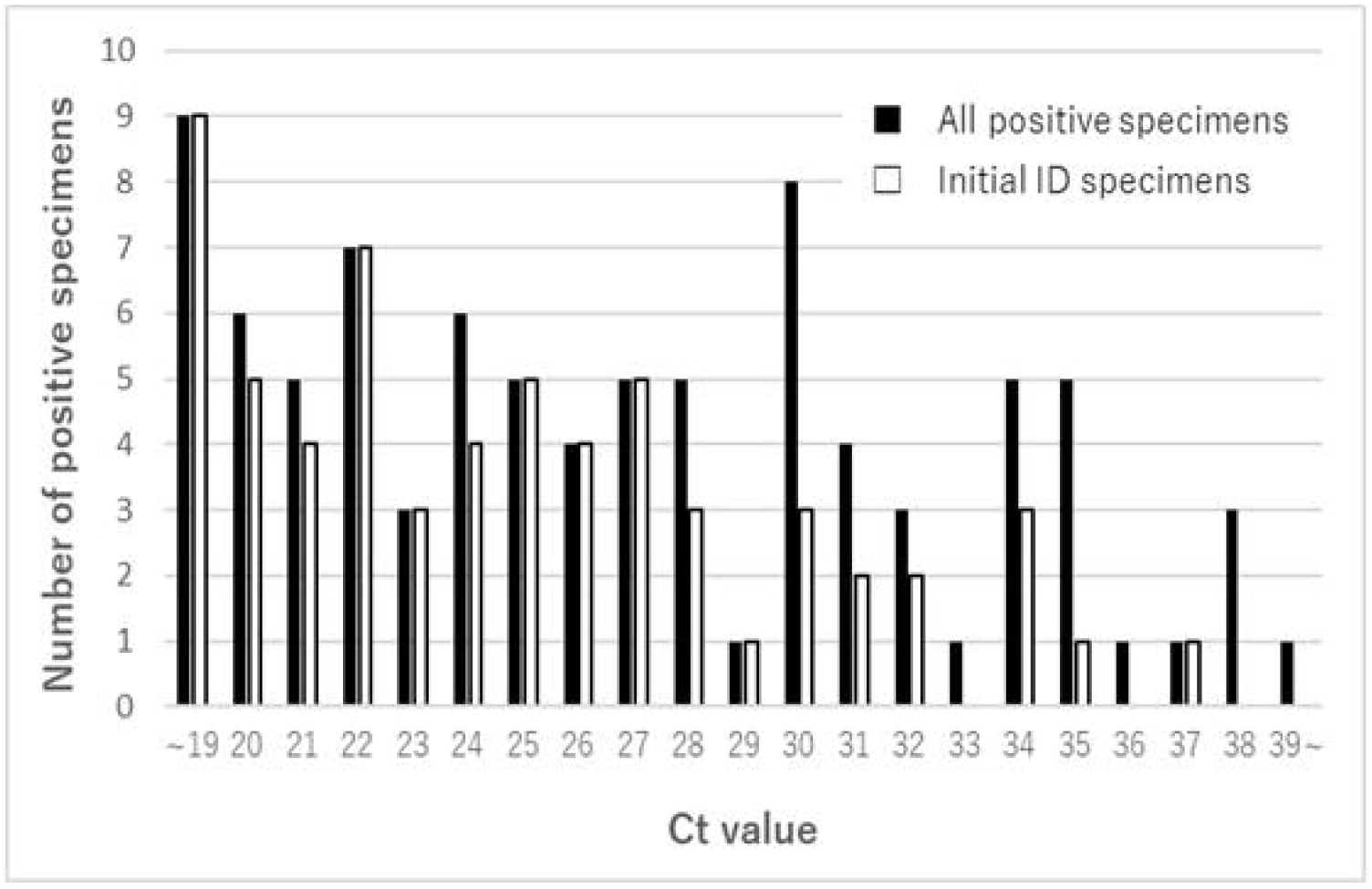
Distribution of the number of positive samples and Ct values Black bar indicates Ct distribution of all positive specimens (N=88), and white bar indicates that of positive specimens with initial ID (N=62).

**Table 1.** The positivity rate in each Ct value range

|  |  | qRT-PCR Ct value range |  |  |  |  |
| --- | --- | --- | --- | --- | --- | --- |
|  |  | <20 | 20 to <25 | 25 to <30 | 30 to ≤40 | Total |
| Number of qRT-PCR positive specimens (N=62) |  | 9 | 23 | 18 | 12 | 62 |
| ESPLINE SARS-CoV-2 | Positive | 9 | 23 | 16 | 2 | 50 |
|  | Negative | 0 | 0 | 2 | 10 | 12 |
| Concordance rate (%) |  | 100.0 | 100.0 | 88.9 | 16.7 | 80.6 |

## Discussion

In this report, we have analyzed the Ct value distribution of the COVID-19 patient specimens received from clinical institutions in April 2020 and evaluated the performance of the rapid antigen testing kit. Initial ID samples likely consist of the specimens collected from patients who were suspected of SARS-CoV-2 infection for the first time, and the other samples probably come from the COVID-19 patients for monitoring purposes and to check for negative conversion. Our analysis has revealed that Initial ID samples exhibited Ct value distribution at low range compared to the other samples. Our results also indicated that, although the NAT method is suitable for monitoring and assessing negative conversion given its higher detection sensitivity, other methods such as antigen rapid testing can effectively detect infected patients at their initial visit. Antigen testing will also provide a way to rapidly screen high-risk populations with sufficient accuracy.

In Japan, rapid antigen testing has been widely used to detect influenza infection, and its clinical usefulness is established ^6^. On the other hand, the clinical value of such tests for COVID-19 has not been investigated, and no evidence was available. In this study, we have revealed that the rapid antigen test correctly detected 80.6% of the positive Initial ID samples as positive, suggesting that it is a useful tool to identify high-risk patients from the population who are suspected to be infected with SARS-CoV-2 for the first time.

PCR testing to detect SARS-CoV-2 is extensively performed globally, but the relationship between Ct value and infectiousness remains unclear. As PCR test is sensitive enough to detect 1 to 5 copies of viral genome in a sample, it may give positive results to patients with extremely low viral load who might do not contribute to virus dissemination, or amplify possible trace of contaminants in specimens. It has been reported that viral isolation and sequence analysis are difficult in specimens with high Ct value^7,8^. Infectiousness of patients who were tested positive with high Ct value, as well as their risks, remain to be clarified.

Rapid antigen testing has several advantages over NAT; it is easy to manipulate, it requires no equipment and investment, and the results can be obtained at the point of care. Our study has revealed that the detection limit of the rapid antigen testing was around Ct=28-30. Ct value distribution data indicated that the rapid antigen tests show approximately 80% positivity in Initial ID samples, suggesting that these tests are useful in distinguishing high-risk patients during an epidemic, and in emergency settings.

NAT is a powerful method, but it is not the only option. Required sensitivity, cost, available medical resources in each region should all be taken into account to shape the optimal testing strategy worldwide to cease COVID-19 pandemic.

## Data Availability

The data that support the findings of this study are available from the corresponding
author upon reasonable request.

## Conflict of interest

SRL Inc. is a subsidiary of Miraca Holdings Inc. Miraca Holdings Inc. holds all stock of Fujirebio Inc.

## Notes

### Competing Interest Statement

The authors have declared no competing interest.

### Funding Statement

No external funding.

### Author Declarations

Declared by the Miraca Institutional Review Boards that this review does not require ethical approval, based on guidelines (Ethical Guidelines for Medical and Health Research Involving Human Subjects) released by the Ministry of Health, Labour and Welfare

